# The use of Reiki in medical settings: personal journeys of Reiki practitioners

**DOI:** 10.1101/2025.11.24.25340938

**Authors:** Sonia Zadro, Peta Stapleton

## Abstract

Reiki is an energy healing or biofield therapy (BT) growing in popularity and developed in Japan at the end of the 19^th^ Century by Mikao Usui of Kyoto (1). It is a non-invasive treatment whereby the practitioner gently places their hands on or close to a client’s body in a sequence of positions that correlate with the major organs and energy systems of the body (2). It is believed energy can be transferred to promote self-healing and restore balance. It is not affiliated with any religion or belief system and is used to complement not replace medical care.

The practice of Reiki is gaining traction in medical settings, with growing evidence it may benefit a range of problems. However, Reiki is an alternative Complementary and Alternative Medicine (CAM) treatment, a Biofield Therapy that is incongruent with the mainstream biomedical paradigm. As such, obstacles to its integration may be expected. This study aimed to explore the subjective experience of Reiki Practitioners (RPs) delivering Reiki in medical settings, using qualitative research methodology of reflexive thematic analysis.

The main topics and themes to emerge were: (1) Positive experiences of practicing Reiki for Reiki practitioners and patients, (11) Negative experiences of practicing Reiki in these settings sometimes due to negative attitudes towards Reiki from other staff and sometimes due to the practical and emotional challenges of delivering Reiki in a medical setting, and (111) Changes needed to promote Reiki in medical settings, in particular training for staff and Reiki practitioners.

It is recommended that when implementing Reiki in a medical setting this is accompanied by an education program using clear consistent language for both staff and Reiki practitioners. This should include information about Reiki research, it’s on-site use and effectiveness, and how to deliver Reiki ethically and consistently while providing support for Reiki practitioners.

## Background and Rationale for Research

### Growing Recognition of CAM and BT

CAM practices, BT, and Reiki are gaining recognition as accepted disciplines internationally. The World Health Organisation (WHO) now advocates for the use of traditional, complementary, and integrative medicine through the *Traditional Medicine Strategy 2014-2023 document,* with a focus on person-centred care and the safe regulation of products and practices (3). Biofield Science is now also viewed as an accepted discipline by some government health organizations, and the term ‘biofield therapies’ was accepted by the U.S. National Institute of Health in 1992 (4). Furthermore, the term ‘biofield’ has been accepted by the U.S. National Library of Medicine as a medical subject search term, the U.S. National Institute of Health (NIH) sponsored research centres for Frontier Medicine in Biofield Science in 2002, (5), and the NCCIH in the U.S. has formally classified ‘energy medicine’ under the broad term of “mind-body practices” (6).

### Reiki’s Growing Efficacy in Treating Clinical Populations

Although Reiki cannot be understood within the established materialistic biomedical paradigm there is growing evidence for its effectiveness. A recent scoping review of Biofield Therapies (BT) found 71 peer reviewed randomised controlled trials (RCT) examining the effectiveness of Reiki with 61 showing positive or some positive significant results and 10 showing non-significant results. No negative results were found (7). In particular, Reiki has demonstrated efficacy with many clinical populations who access inpatient and outpatient medical care. There is growing evidence of the effectiveness for Reiki over placebo for clinically relevant symptoms of anxiety, depression and stress, moderate to high blood pressure, acute and chronic pain, and chronic conditions (8, 9). Additional RCTs have also found Reiki to produce significant positive outcomes other conditions requiring hospitalisation. These include Reiki for cancer patients (10–16), palliative care (14, 17), pain (16–21) cardiac disease (22) pulmonary disease (23), mothers of hospitalised children (24), third molar surgery (21), and healthcare professionals (25, 26). As such, Reiki may provide a valuable contribution to a variety of medical settings.

### How Reiki is integrated into Mainstream Medical Centres

Reiki is integrated into conventional medical care in three ways. In some institutions, medical staff learn first-degree Reiki and use it for self-care. In other hospitals, RPs offer treatments to patients and staff, and in other hospitals, education programs are training patients, family members, and caregivers first-degree Reiki (27). Reiki has been provided as an option throughout acute care inpatient settings, including endoscopy (28, 29), oncology infusion units (30–32), perioperative units (33–35), hospice care settings, emergency rooms, psychiatric settings, operating rooms, nursing homes, paediatric, rehabilitation, obstetrics, gynaecology, and neonatal care units, HIV/ AIDS, organ transplantation care units (27) and operating theatres (34).

### Reiki’s use in Medical Centres Internationally

Internationally, there has been a growing acceptance of energy healing and Reiki. UK healers have been working in UK hospitals since 1959, and the UK General Medical Council first allowed doctors to refer patients to healers in 1977 (36). In one UK study (36), a questionnaire survey was sent out to 127 health centers across the UK in 2008-2009 which offered healing services. Sixty-seven questionnaires were returned from 38 conventional cancer care settings, of which 16 were based on the National Health Service (publicly funded). These included 25 hospices, eight hospitals, and five Macmillan centers. Of these, 33 centers offered Reiki. Overall, about half the healers communicated with the patient’s doctor, and most healers believed it to be very well accepted, with most reporting that their non-healer colleagues perceived it as useful or, usually, very useful. Feedback on these services was highly positive; however, a third of healers had no administrative support, and 40% were unpaid, which was a perceived difficulty (36)

In the United States, a 2012 survey (37) also found that over 55% of Integrative Medicine clinics across the U.S. used healing touch or Reiki to assist in treating a variety of problems, including cancer, chronic pain, immune disorders, heart disease and arthritis, gastrointestinal problems, diabetes and allergies, and obesity.

In the U.S, mainstream hospitals including Reiki as part of their program or offering it as a service include the following hospitals and a number of these are highly ranked including the integrative oncology centre at The Mayo Clinic (31) which has been ranked as the number 1 hospital in the world (38), The Cleveland Clinic, Ohio (39), which has been ranked as the number 2 hospital in the world (38), and the New York Presbyterian Hospital, New York (40) which has been ranked as the 14th best hospital in the U.S. (38).

To date, there have been no studies exploring or assessing the use of Reiki in the Australian population or in Australian medical centres. As such, it is not easy to determine to what extent it is being accessed or integrated in Australia.

### Obstacles to Reiki’s integration in Medical Settings

#### Personal Resistance

There are personal, paradigmatic, and pragmatic obstacles to Reiki’s mainstream integration in these settings. In an unpublished qualitative study (41), fifteen licenced mental health professionals (LMHP) who found Reiki a valuable tool in therapy were interviewed about their use of Reiki. Their concerns included the following themes:

(1) Concerns about whether Reiki was viewed unscientific or a fake science by other LMHP or clients, versus wanting to use Reiki.
(11) Wanting to find a like-minded community of professionals who were certified in Reiki to discuss its application.
(111) The need for psychoeducation about Reiki research for LMHP and clients.
(IV) A need to know that clients were receptive to the idea of Reiki before it was introduced.
(VI) A fear of violating ethical codes related to Reiki not being accepted as a tool by authorities.

#### Paradigmatic Resistance

Perhaps the main resistance to Reiki and BT in general involves the notion that energy therapies suggest the existence of a life force behind consciousness, historically referred to as ‘vitalism.’ In this way, energy healing is suggestive of Cartesian dualism – the idea that we have a spirit as well as a physical body. Science has always firmly rejected such a notion. As such, this kind of paradigmatic resistance may persist for some time unless, either the causal factors for Reiki and the biofield are clearly explained in ways that do not incorporate these ideas, or mainstream science becomes more open to such possibilities, the latter of which presently seems unlikely.

#### Pragmatic Obstacles to the integration of BT

There are also pragmatic obstacles to integrating BT. Some of these are outlined in a white paper published in Global Advances in Integrative Medicine and Health (6) which included the following: A perceived need for uniform agreement on accepted terminology and definitions to be used in Biofield Science, a clearly defined BF mechanism, standardized technology for BF assessment, and unified standards of practice. This lack of uniformity and standardization was seen as contributing to confusion amongst the medical community, educators, and patients. There was also a lack of common educational standards, training, and professionally accredited modalities, as well as a lack of research and funding for Biofield Science in general.

## Research Objectives

Given the personal, paradigmatic and pragmatic obstacles outlined above, this study aimed to explore Reiki Practitioners’ (RPs) experiences of practicing Reiki in mainstream medical settings to identify whether they encountered similar and/or other kinds of obstacles, and to identify factors which may aid its integration to assist both RPs and other medical staff.

## Method

### Study Design

Because this was an exploratory study focusing on subjective experiences within an emerging, alternative area of integrative medicine, a qualitative methodology was chosen – reflexive thematic analysis (reflexive TA). This methodology focuses on subjective experience to identify and report patterns or themes within response sets and interprets them in light of the research questions. It is not aligned with any pre-existing theoretical framework and may reflect reality or explore what lies beneath the surface of reality (42). Reflexive TA also allows for insight into potentially unexpected themes in a new area. Given this, along with its subjective focus and flexibility, it was viewed as an appropriate methodology for a novel treatment and emerging topic in healthcare under study. The researcher was a clinical psychologist with 30 years’ experience conducting this research as part of her PhD in Reiki and Biofield Science at Bond University Australia. Guidelines of *Consolidated Criteria for Reporting Qualitative Research* was used as a guide (43). Interviews were recorded, transcribed, and stored on file with TEAMS software, and demographic data was collected through a Qualtrics survey. A semi-structured interview using 13 questions allowed participants to freely express their thoughts and feelings, while allowing for consistency in the topics under discussion. Interviews lasted between 30 and 50 minutes.

### Ethics Statement

Ethical approval was granted by the Bond University Human Research Ethics Committee (BUHREC: Number SZ00084) and the U.S.-based ethics committee, the National Institute for Integrative Healthcare (approval number NIIH20250201). All participants provided written consent prior to their participation in the research.

### Participants

Eight Reiki practitioners (RP) were interviewed, two males and six females. Four of these were from Australia, and four were from the U.S. This number was considered sufficient for a preliminary exploration of the topic under study as the requirement of data saturation is deemed inconsistent with the assumptions and values of reflexive thematic analysis (44). Participants were recruited through an advertisement on the Reiki Australia website, or word of mouth suggestions from Reiki Australia and The Centre for Reiki Research. U.S participants were included because of the limited number of medical settings that use RP in Australia and because medically based Reiki programs are more widely established in the U.S. It was thought that RP from the U.S. may therefore serve as a potentially informative comparison to Australian RP perspectives. Recruitment took place between the 17^th^ December 2024 and the 7^th^ February for Australian participants, and between 2^nd^ and 30^th^ February 2025 for U.S participants.

Inclusion criteria for RP participants were: a minimum age of 18 years who had practiced Reiki on patients in a medical setting. There was no minimum level of experience for RP. There was no prior relationship between the researcher and any participant at the commencement of the study. Interested participants responding to the advertisement contacted the researcher by email. Word of mouth suggestions were emailed a brief invitation to participate and all accepted the invitation. All participants were then sent a letter of invitation outlining the research aims and informed consent assuring them of confidentiality, which they signed and emailed back with a preferred interview time. All participants were informed of their right to withdraw consent at any time. All eight participants who signed up for the study completed an interview with no attrition.

Medical settings where the RP worked mainly included two large university hospitals, a mental health clinic, and two palliative care hospices. Most centres or units were for cancer patients or palliative care patients. Sometimes, the RP had worked at more than one location. Most patients were treated for cancer related symptoms of pain, anxiety, stress, nausea, and other issues such as small bowel obstructions or phantom limb pain. Some also experienced pain or anxiety related to end of life. In some settings, Reiki was also offered to staff and family members of patients upon request. Sessions ranged from approximately 10 to 60 minutes, although this was usually flexible and sometimes based on patient need and the degree of symptom relief experienced. RP’s years of experience in providing Reiki in medical settings ranged from under 3-months to more than 20 years. All were unpaid for their practice of Reiki in these settings except two RP who were paid as Reiki Program Co-ordinators. Four RP were also registered health professionals, and one was a pastoral care provider. See Table 1 for participant characteristics.

**Table 1:** Participant Characteristics.

|  | P1 | P2 | P3 | P4 | P5 | P6 | P7 | P8 |
| --- | --- | --- | --- | --- | --- | --- | --- | --- |
| **Country** | U.S. | Australia | Australia | Australia | Australia | U.S. | U.S. | U.S. |
| **Gender** | Female | Female | Female | Female | Female | Female | Male | Male |
| **Medical Setting** | Mental health clinic.<br>Inpatient Oncology Unit. | Regional Integrative Cancer Hospital | Palliative Hospice. Private hospital inpatient and outpatients. | Regional cancer centre | Palliative Care Hospice.<br>Inpatient Hospital | University Inpatient and outpatient Hospital | Acute care state hospital – inpatient and outpatient.<br>Integrative Oncology setting. | University Hospital Oncology Centre inpatients |
| **Length of time Reiki program running** | Formal Reiki program several years. | No formal Reiki program running but Reiki offered for several years. | No formal Reiki program but Reiki had been offered for approximately 30 years. | No formal Reiki program. Reiki not offered prior to her service. | No formal Reiki program but Reiki offered for several years. | Reiki program officially running 3 years, unofficially Reiki offered several years before this. | Reiki program officially running 8 years.<br>Reiki provided unofficially 17 years prior to this by one chaplain. | Undisclosed |
| **Conditions Treated** | Mental health issues<br>Mainly oncology patients. | Oncology inpatients and outpatients | Mostly palliative care patients | Cancer patients | Palliative Care patients | Adult Oncology patients, Mother-Baby, labour and delivery and pre-natal inpatients. | Cancer patients (pain, nausea, anxiety), small bowel obstruction, phantom limb pain, burn survivors (2<sup>nd</sup> degree burns). Also given to staff and patient's families. Reiki provided to 250 nurses last year, and approximately 2000 sessions / year overall | Cancer related symptoms e.g. fatigue, pain, stress |
| **Length R session (approx)** | Inpatients: 30 mins.<br>Outpatients: 20 mins | 40 mins | 45 mins | 30-45 mins | 30-60 mins | 15-20 mins | Needs basis until symptoms reduce significantly. 10-60 mins usually | Undisclosed |
| **Years practicing R in medical setting** | >7 years | 3 months | 18 months | 3 months | >2 years | >20 years | >12 years | < 3months |
| **Registered healthcare professional** | Undisclosed | Yes | Yes | No | No | Yes | No | Yes |
| **Volunteer / Paid** | Volunteer | Volunteer | Paid as an Outpatient Co-ordinator | Volunteer | Volunteer | Paid as a Reiki Co-ordinator | Paid as a Reiki Co-ordinator, 2 other RP paid and 3 other RP unpaid | Volunteer (paid medical specialist) |

### Data Analysis

Interview responses were analysed through thematic analysis of transcribed interviews using NVIVO software. Support themes with quotes were provided from transcriptions. Interview audio recordings, transcriptions, and analysis were stored in an approved Bond University data storage site. De-identified data (or any publications) were stored in an OFS.IO weblink. The six stages of reflexive TA were applied to the transcripts (42, 45). Interview transcripts were coded as a meta-synthesis by one PhD candidate (the lead author).

For Stage 1: *data familiarisation*, the researcher listened to all interviews, corrected transcribed words, and de-identified references to the participant. Transcripts were grouped into responses to interview questions to deepen familiarity. For Stage 2, generating *initial codes* involved reviewing responses to questions and creating a list of ideas or codes identified in the data and assigning labels to them. During Stage 3: *Generating initial themes*, original transcripts were re-read, and codes were grouped into broad themes and subthemes. During Stage 4: *Developing and reviewing themes*, codes were checked to ensure they fit with each theme or subtheme. Themes were re-assessed to ensure they best captured the main shared ideas expressed in the data. During Stage 5, *refining and naming themes*, themes were further streamlined and codes were regrouped to better fit consistent themes ensuring each theme/ subtheme was a unique idea. Finally, writing the report required an overall interpretation and reporting of findings to form a meaningful narrative with clear conclusions.

While some researchers assume it is important to control researcher bias in thematic TA (46–48), other leading researchers in the field have argued that quality coding does not require more than one coder or collaboration because subjectivity is seen as a strength in reflexive TA, not as something to be controlled. Rather, it has been argued that reflexivity and interpretive depth account for good analysis (44, 49, 50). As such having only one coder was viewed as acceptable. Reflexivity, creativity and the acknowledgement of the author’s generative role in the research is also recommended (49). In light of this, the primary researcher maintained reflexive journaling and ongoing discussions with her supervisor to maximise reflexivity, creativity and promote awareness of how her personal beliefs might subjectively impact the process of identifying themes. The author had practiced Reiki but had never done so professionally and self-reflection on how this impacted her interactions with participants and her analysis was discussed.

## Results

Reflexive thematic analysis produced three topic areas with related themes. The three topic areas were *Positive experiences of Reiki in medical settings*, *Negative experiences of Reiki in medical settings*, and *Changes Needed to Promote Reiki in Medical Settings.* See Table 2 below for a summary of Topics, themes, and related subthemes.

**Table 2:** Reflexive Thematic Analysis: Topics, Themes, and Subthemes for Reiki Practitioners Experience of Delivering Reiki in Medical Settings.

| Topics and Related Themes | Subthemes |
| --- | --- |
| Topic 1: Positive Experiences of Practicing Reiki in a Medical Setting. |  |
| <u>Themes</u><br>1:1 Positive Experiences of delivering Reiki for the RP and patient.<br>1:2 Reporting Positive Outcomes of Reiki<br>1:3 Feeling Supported by Management and Staff | (a) Reiki as deeply rewarding for the RP and patient<br>(b) Receiving while giving Reiki<br>(c) Giving to the patient, not taking |
| **Topic 2: Negative Experiences of practicing Reiki in a Medical Setting**<br><u>Themes</u><br>2:1 Negative attitudes towards Reiki<br>2:2 Challenges of a Medical Setting | (a) Suppression and stigma<br>(b) Feeling unvalued<br>(c) Religious or world view objection/ general doubts about Reiki<br>(d) Lack of awareness about Reiki and it being offered<br>(a) Emotional demands<br>(b) Limited physical access to patients<br>(c) Time constraints with competing appointments<br>(d) High Demand / Limited availability of RP |
| **Topic 3: Changes Needed to Promote Reiki in Medical Settings** |  |

| <u>Themes</u> |  |
| --- | --- |
| 3:1 Staff training for Reiki in Medical Settings | (a) Reiki research on efficacy and how it works<br>(b) Evidence of on-site effectiveness<br>(c) The importance of direct experience of Reiki for staff<br>(d) Clear consistent explanations of Reiki in their language<br>(e) Encouraging referrals and volunteers from staff |
| 3:2 Reiki Practitioner (RP) Training for Medical Settings | (a) Prepare RP for the medical setting<br>(b) How to administer Reiki in a medical setting<br>(c) Uniform, consistent practice and explanations<br>(d) Support for RP |
| 3:3 Ethical considerations for Staff and RP | (a) Reiki does not replace medical care, is non-religious and is not used to diagnose<br>(b) Always offering a choice for Reiki and touch<br>(c) Governance, professional membership |
| 3:4 Awareness and Promotion of Reiki | (a) Awareness and promotion of Reiki research<br>(b) Management and staff advocacy and support for Reiki<br>(c) Providing education about Reiki internally and externally<br>(d) Teaching Reiki to staff and patients<br>(e) Using accessible referrers and appropriate volunteers<br>(f) Unified consistent understanding and explanations of Reiki and Biofield Science<br>(g) Disconnect between intuitive self/healer and researcher/medical practitioner |

### Topic 1: Positive Experiences of Practicing Reiki in a Medical Setting

#### Theme 1:1 Positive experiences of delivering Reiki for the RP and patient

##### (a) Reiki as deeply rewarding for the RP and patient

All participants spoke of how providing Reiki in a medical setting was a positive experience, most describing it as “beautiful” and rewarding for both them and the client. Most participants felt very strongly about these positive experiences, expressing, in particular, how Reiki provided the client with a sense of deep peace, relaxation, and often pain reduction.

> *“Most of the time they’re relaxed and they fall asleep and sometimes they wake up and then they were just very, very, very appreciative, and they say ‘Ohh, I’ve never experienced anything like this before, a deep, deep peace’, and they relax.. After I give Reiki, they can just sleep. They sleep for a long time.”* (P5)

> *“Oh I had a guy they wanted me to go see. He said he hasn’t slept in three days. So I went to see him and he said, ‘This pain is awful.’ He can’t sleep, he’s writhing around a whole nine yards, and I explain what I’m gonna do, and I start Reiki and he settles right down and he says, ‘Oh my God, I haven’t been without pain for three years.”* (P7)

> *“I had one experience with a patient who was there for end-of-life care and I did a Reiki session with her and she just felt so joyful, and at the end of the session she’s like, ‘Thank you… I just feel so happy. I feel so content.’ I … reflected on that a lot after that session, for probably weeks and months afterwards, and I just thought how nice it must be to have somebody, stop to acknowledge that even though you’re at the end of your life, that at that moment you were really, really happy.”* (P3)

There were also numerous examples of pain relief for problems such as multiple brain myeloma, pain post-surgery after the removal of a brain tumour, phantom limb pain, and small bowel obstruction.

> *“Somebody’s got a small bowel obstruction and its very, very painful, and they’re gonna go through an operation, and we do Reiki for them, and it clears. Wow! You know, I mean and the doctors are flabbergasted, so I mean, it’s hard to explain. It’s just like – I love what I do.”* (P7)

##### (b) Receiving while giving Reiki

Along with describing the experience of giving Reiki as beautiful or positive, some RP spoke of how, when giving Reiki, they also received it. They described it as fulfilling because they felt the energy moving through them as well.

> *“People are like, ‘Wow, you know that time together was just profound.’ So I find it changes my life every time I give it and I come out better. We both come out of the room better then we went in… I say, you know, there’s only a few things you can do where you are giving and receiving at the same time. And this is one of them.”* (P6)

##### (c) Giving to the patient, not taking from them

Another topic several participants spoke of was how they believed patients appreciated that the Reiki practitioner was giving something to them rather than taking something from them. They spoke of how patients often felt a sense of medical trauma or their choices taken away when given a diagnosis. They may have also felt a lack of personal interaction with treating health professionals and that, in comparison, Reiki was something they could choose, and during a treatment, they valued receiving nurturing and a sense of connection to the RP.

> *“So if you think about somebody who’s laying in a hospital bed. A lot of people who come in and see them were staff placed and are there to take something from them. Whether it’s taking a blood test, the blood pressure, asking about their bowel movements.. it’s all sort of, ‘I want this from you, I want this from you, I want this from you’… I think for someone like a RP to go in and give something back is really beautiful.”* (P3)

#### Theme 1:2 Reporting positive outcomes of Reiki

All RP felt that Reiki added value to the patient’s well-being and health outcomes. Nearly all said this was based on patient feedback and spoke enthusiastically about this. Two mentioned that while Reiki nearly always added value, occasionally, a patient did not feel much from the session or did not esonate with it. Most RP, however, spoke of patients returning for more Reiki and being unable to meet patient demands.

> *“I actually was surprised I was booked out from the word go because I thought this is something I’m going to have to really promote”* (P3)

> *“So we have documentation now and there’s tonnes of it now of what the positive results have been. It’s phenomenal where it’s gone.”* (P7)

#### Theme 1:3 Feeling Supported by Management and Staff

Some RP said they generally experienced no resistance from staff or management. This included three RP who worked in large medical settings in the US where they had practiced for a long time in well-established programs. Two Australian RP also experienced minimal resistance and felt generally supported by management, again in settings where providing Reiki was more established. This appreciation was directly expressed to the RP or was evident in the staff’s extra steps to accommodate the RP’s needs. In one Australian hospital, a separate quiet room with a massage table had been set up primarily for RPs to provide Reiki to patients and families. While at another Australian hospital a RP spoke of how the nurses were happy to quietly turn the IV drip alarms off before they ‘buzzed’ so the alarms did not disturb the tranquillity of the session.

> *“Oh they were super, super-responsive and really appreciative and supportive of me being there which is yeah, that’s what you want.”* (P3)

In a large US hospital where a Reiki volunteer program had been established for many years, Reiki had been funded through philanthropy. This RP reported having no pushback and no negative feedback from any staff or management in all the time she had been there, as well as the full support of volunteer leadership, and the board leadership team. Another US RP from a different hospital said that the chief operating officer was so supportive of Reiki volunteers she told the RP he needed to pay the RP volunteers.

> *“You need to pay people, cause you’re gonna lose them.”* (P7).

### Topic 2: Negative Experiences of Practicing Reiki in a Medical Setting

#### Theme 2:1 Negative Attitudes towards Reiki

Despite the deeply positive experience of delivering Reiki by most RPs, and the support most of them felt, most RPs also had some negative experiences of Reiki in medical settings. Some felt unvalued for their service, a sense of stigma and suppression attached to providing Reiki, and they encountered religious and worldview objections to Reiki, or a general sense of resistance towards Reiki practice. This was also evident when staff had been primarily supportive on other occasions.

##### (a) Suppression and Stigma

Most RP at times felt negative attitudes directed towards them for practicing Reiki or a general stigma towards their more spiritual worldview. One RP, a specialist physician, described the reaction of other members of his profession to Reiki.

> *“There’s a lot of suppression. I’ve given a lot of talks… I’m pretty open about it and I can feel people’s snicker. I’ve talked about this at medical conferences and I can tell that people don’t really, you know, respect it…I’ve been told by my mentors not to do research in this because it’s alternative.”* (P8)

This RP went on to describe how he felt this suppression from others in his profession, relating not only to Reiki but in relation to spiritual experiences in general.

> *“I find it very similar to conversations about religious experiences… There are many people who if you open up a conversation, (people have) had experiences they would never tell in public… It’s great to be scientific, but there’s a suppression of lived experience.. I can tell people I’ve had all these experiences and they’ll tell me, ‘No you haven’t,’ or…say ‘Well that doesn’t make any sense.’ Like well, why don’t you spend 10,000 hours meditating and learning this and then we’ll have a conversation. They don’t want to go through that, but they have a strong opinion because this doesn’t make sense to them.”* (P8)

Another RP spoke about how she was not permitted to speak about Reiki or promote it, and that she did not understand why, considering she was only there to help the patients.

> *“You were only allowed to talk about it if someone approached you but you weren’t allowed to hand pamphlets out to people…I couldn’t understand why when I was there to help, why it was limited on the way I could express and talk about Reiki… I’m not here to convince people the value of Reiki. It almost felt like I had to do a hard sell… there was a lot of resistance.”* (P2)

Another RP spoke of personal interactions with staff members who were sceptical about Reiki.

> *“There’s a nurse here that says … ‘I don’t believe in that crap’..and that’s okay, you know it’s a personal thing… And we had a doctor, ten years ago who used to call it Reiki Fake-i, and he would say you know, ‘It’s all mind over matter.’ And I would say, ‘So what…if it is and it works what do you care?’ So we have enough documentation now and there’s tonnes of it now, of what the positive results have been.”* (P7)

##### (b) Feeling Unvalued

Negative attitudes also came in the form of feeling unvalued as a RP or that complementary therapies generally were not valued enough. This often took the form of an unwillingness to remunerate a RP in any way, deriding comments, or cutting services due to misinformation. One Australian RP (P2) was told if she volunteered weekly for a few months they would advertise her services. She said this did not happen. She was also told she could negotiate a fee after several months but that this got “shut down”. Finally, she asked if they could pay for her parking as they had paid for another CAM practitioner’s parking. This was also refused.

> *“So that made me feel that the value wasn’t there. You value me so much but you’re not willing to pay for parking for me? For volunteering three and a half hours of my time?… That was why, yeah, you can understand my perception of why there’s a lot of barriers there.”* (P2)

Another volunteer overheard some staff laughing about a patient because of her esoteric beliefs and feeling devalued by their comments.

> *“They just scoffed and laughed and you know, I was sitting there quietly going, ‘But it works and I understand and… why are you devaluing something that’s very intrinsic to me?’ … I guess hearing all those sorts of comments on the wards over the years, it takes a bit of courage to sort of step out of that box and say, hey this is what I do and this is why it’s so great.”* (P3)

Another volunteer participant (P4) felt unvalued when asked to leave based on misinformation about Reiki and touch.

> *“Meanwhile yoga’s allowed to be there and (has) a paid teacher…But I’m not going to fight the fight because I’m a volunteer. I shouldn’t, you know. It’s like swimming upstream.”* (P4)

##### (b) Religious or Worldview objections and general doubts about Reiki

Some participants said that some patients had a religious, faith-based or worldview objection to Reiki. These were often from the Christian faith.

> *“There’s quite a bit of resistance from people with… mainly strong Christian backgrounds and that it’s against the religion. If management can …explain better that Reiki is non-religious. Because they believe that, oh, you know, ‘That’s the spirit of the devil.’”* (P5)

Another participant noted that, although Reiki was popular in India, many gurus opposed it. He said this affected his personal journey of accepting and using Reiki as a treatment.

> *“It’s not in sync with most people’s belief systems …(then) there are people on the other side of the spectrum. There are many gurus in India, for example, who are very staunchly again using Reiki because they have a different take on it….It was enough to make me question whether I should do this…(but) for me I think if it’s gonna help someone I’m not really worried about the, you know spiritual, existential side of it and I’m just trying to help someone feel better. So I stay within that focus and then I strongly believe that it helps and have felt it myself.”* (P8).

##### (c) Lack of awareness about Reiki

Sometimes, negative attitudes related to a lack of awareness that Reiki was being offered, what it was, or what it involved. This appeared to come from a lack of education and/or promotion of Reiki. It was generally evident in medical settings where Reiki was less established.

> *“It kind of flies very low under the radar… it’s not talked about a lot.”* (P3)

> *“It was more so, doctors and nurses just weren’t aware of Reiki. They thought it was woo-woo…”* (P2)

> *“Our biggest obstacle was education. Most people weren’t sure what we were doing.”* (P7)

Another participant was asked to leave on the incorrect basis that Reiki involved touch. There was a lack of awareness that Reiki could be delivered near the body without touching it.

> *“I was a bit taken aback by this decision considering it was going so well… to have these people who I don’t know, ghosts in the cupboard… who have made this decision out of ignorance because you can also provide Reiki without even touching - hands above.”* (P4).

#### Theme 2:2 Challenges of a medical setting

Another area that created challenges for RP was the demands of the medical setting – both emotionally and physically, in terms of the medical environment, and the time restrictions often imposed by competing appointments. Finding enough RP volunteers to meet patient demand was also often difficult, as RPs were generally unpaid.

##### (a) Emotional demands

Some participants stated that new RPs needed to be well-prepared for a medical environment, as it differed significantly from a wellness clinic or working from home. Patients could be acutely unwell, dying, physically very damaged, and immobile.

> *“You’re seeing some … pretty crazy stuff, you know, people all broken.. Again the biggest drawback is there may be things you don’t want to see. We have a burn clinic that can be pretty intense. We had a girl who lost her son to a stroke. She said she was okay, but she went in the room and it was overwhelming. She was right to come back out.”* (P7)

##### (b) Limited physical access to patients

Most RP said they felt limited by the physical environment of a medical setting in gaining easy access to the patient but learnt to work around it.

> *“It was difficult in so far as you’re working around a hospital bed with, you know drips and tubes and all kinds of other things. You can’t get all the way around the bed, and of course the bed is much lower than a massage table..”* (P1)

##### (c) Time constraints with competing appointments

The medical setting also imposed numerous demands on patients, including staff checks, monitoring, various allied health and medical assessments, as well as interruptions from the patient’s visitors. This meant the RP needed to be much more flexible than when working in a private clinic or Wellness Centre.

> *“So, sometimes your Reiki session will get disrupted because they need to come and do their medication rounds or you know the physio needs to do their review, and I say okay, this is important too… You know the nurses themselves are very busy… they just need to get the meds and get through and answer the call bells and things like that.”* (P3)

##### (d) High demand / limited availability of RP in medical settings

Because medical establishments are usually not prepared to pay for RP given its alternative and emerging status, it is not always easy to find RPs willing to volunteer. Most participants in this study were unpaid volunteers and the demand for Reiki treatments often exceeded supply.

> *“It’s phenomenal where it’s gone. I mean, we’re in the 2000 range for sessions a year. I don’t have enough people to do the sessions that we need to do.”* (P7)

### Topic 3: Changes Needed to Promote Reiki

All RP mentioned or spoke at length about the need for staff training to promote awareness and understanding of Reiki, the need for adequate RP training to work in medical settings, and the need to promote more awareness of Reiki generally. Ethical issues were also frequently raised as they were considered an important part of training for both RP and staff.

#### Theme 3:1 Staff Training for Reiki in Medical Settings

##### (a) Reiki research on efficacy and how it works

All participants emphasized the need to train staff on the current status of Reiki research, its efficacy, and research related to its mechanisms of action, specifically its impact on stress, blood pressure, and the autonomic nervous system.

> *“There just needs to be greater awareness… having more research out there which makes it that little bit more palatable to medical teams because they’re not interested in the woo-woo stuff. They like the facts and the figures and evidence, right?…If medical people can accept that we have a biofield, which we do, we’ve got a magnetic field from the heart, we’ve got that electrical field from our brain, then if they can see we can make interventions in that sphere that can reduce things in the physical body then I think it would be a lot easier for them to go, ‘Okay you’re doing something.’”* (P3)

> *“We can’t measure specifically the Reiki energy, but we can measure outcomes, and I think that’s important if our outcomes have been significant.”* (P6)

##### (b) Evidence of on-site effectiveness

Part of promoting awareness of Reiki’s efficacy involved providing patient feedback to medical and other staff, documenting how Reiki felt to the patient and the degree to which it provided symptom relief. Some participants spoke of how this documentation was integrated transparently on the ward so that all treating practitioners could see that Reiki was being used and how effective it had been. One RP felt very strongly about this. He said the effects of Reiki and comments about the treatment are charted with every patient and explained that these stay on the patient’s record for life.

> *“I’ve been adamant that we put the information in a patient’s chart. So I had an orthopaedic surgeon that asked me to go see a patient. He should know what I did and what the results were. So, what sells it is: ‘your patient had pain of ten and it went down to four.’ Now he’s sold. That’s very, very key…In the medical profession we’re talking about science, we’re also talking about metrics. You’ve got to have both. When I tell you of a 65% reduction in pain – that gets noticed to a physician and that’s gonna get noticed to a nurse.”* (P7)

##### (c) The importance of staff directly experiencing Reiki

While providing evidence of Reiki’s efficacy was considered important, half the participants also emphasized the importance of staff and occasionally family members needing to experience Reiki directly to understand what it felt like and how it could help. Sometimes, this took the form of demonstrations as part of educational sessions for nurses. At other times, Reiki was offered to a nurse during a shift or when a nurse appeared to need it emotionally. One participant also emphasized the importance of management or the *“people making decisions*” (P4) needing to experience Reiki firsthand.

> *“You can talk about it all you want (but) I think people need to experience it. So when I’m introducing it, I also demonstrate it, and I think when someone can say ‘Wow,’ you know, and have people witness that… and go ‘Ohh my goodness’… and it’s not just the patient… getting it, I think that makes a huge difference… then people kind of see how it works, even if it’s, you know, oftentimes hard to describe…”* (P6)

> *“We did probably 215 nurses last year… which again sells it… I think the mistake that people make is they’re trying to cultivate it from the outside and convince someone what it is, and you don’t know what it is until you’ve experienced it… When somebody does Reiki for you, you’re sold but you can’t verbally explain it.”* (P7).

##### (d) Clear, consistent explanations of Reiki

Several participants also emphasized the importance of using simple, clear, and consistent language when explaining Reiki and its workings to staff and patients. Several RP felt that everyone needed to agree on the kinds of explanations that were provided, and when speaking to medical staff to use the kind of language they could relate to.

> *“You need to have the right person leading the Reiki program, who knows how to speak to both medical professionals and patients and people who aren’t familiar with it, and to express what it is without putting people off…so it doesn’t scare people.. We have a common language that we can use and that’s integrative…You have to not be too technical and medical, or too what people call ‘woo-woo’ you know, but to have enough knowledge… to have definite processes… I think that’s why our program is successful and growing…”* (P6).

##### (e) Encouraging referrals and volunteers from amongst staff

Participants generally advocated that anyone well-trained and experienced in Reiki, preferably with hospital training, could deliver Reiki in a medical setting. However, certain members of staff were viewed as good potential referrers to RP, such as pastoral care and allied health. Some participants believed that staff working in these professions better understood the emotional needs of the patients because they spent more time talking to them. Another participant strongly advocated for Reiki to be administered internally by staff already working in the hospital, particularly nurses.

#### Theme 3:2 Reiki Practitioner Training for Medical Settings

##### (a) Preparing RP for a medical setting

Most participants emphasized the importance of being prepared for a medical environment and the necessity of having proper training before they began delivering Reiki in this setting. Some had previously worked in hospitals in another professional capacity. Most were required to undergo first aid training, hospital orientations, and police checks, and were given an induction day to prepare them; however, unless they volunteered in a centre with an established Reiki program, no training tailored specifically to Reiki was provided. In some more established Reiki programs, RPs were required to shadow an existing volunteer until they felt ready to deliver Reiki themselves. One RP also suggested training RP in medical terminology may be helpful.

> *“So they usually shadow me for two or three weeks and then we ask them for at least four hours a week.”* (P7)

> *“Only people who are licensed and who have had experience on inpatient are doing inpatient at this point because we want to make sure you know, leadership and patients and everyone feels comfortable.”* (P6)

This RP also believed that patients benefited from shorter sessions and as the Reiki Co-ordinator, she recommended this.

> *“A lot of times medically challenged people can tire easily so I usually start with like 15 to 20 minutes. If they can tolerate longer, they let me know. Some people stop you before that because it is a lot to have someone in your intimate treatment space.”* (P6)

##### (b) How to administer Reiki in a medical setting

The logistics of how to refer to RPs and how to conduct sessions in medical settings was discussed by some participants. Most participants worked in a setting where a separate person handled the bookings, and bookings were made when a patient requested a Reiki session. The volunteers were then contacted to determine who was available during a specific time period. In some settings, the RP was also a nurse working on the ward and so, the Reiki session would need to fit in with their schedule. In other settings, patients were visited by the RP and directly asked if they would like to experience Reiki. At one medical setting, patients were offered a set number of complimentary supportive treatments over a specified period, and Reiki was included in the list of treatments provided.

In terms of how the Reiki was delivered, several participants spoke of the need to keep this relatively uniform and agreed upon across all RP volunteers with some flexibility. Sessions lasted between 10 to 60 minutes across all medical settings, with most ranging between 20 to 45 minutes per session. In some settings, hand positions were completely flexible, while in others, there were set protocols for hand positions that still allowed the RP to use their intuition to determine where the Reiki was most needed. Another RP said they usually started with around ten minutes but generally continued until the symptoms had significantly reduced or the patient had fallen asleep, which was common as Reiki is generally reported to induce a state of relaxation. Falling asleep was also a sign the patient’s pain had likely reduced.

##### (c) Maintaining uniform and consistent practice and explanations

As with explanations to staff, several participants emphasized the need to provide simple, consistent explanations to patients and uniform practice in general.

> *“..we did kind of have to establish some practice guidelines on, you know how we described Reiki, how we talked to the patient, what kind of language we would use… to make sure we’re all on the same page… to go over consistent practice guidelines. We ended up um having to …bring everybody into a meeting and say, ‘Okay we don’t want to talk about this, we don’t want to talk about that, we want to keep it… just regular Reiki practise and that’s it.”* (P1)

> *“We have a standard operating process… we have an intro script so we’re all saying about the same thing. No one’s going off on their own and adding things that don’t belong… I’ve talked to volunteers and if they’re gonna not follow the rules they can’t volunteer with us because you don’t bring in things that aren’t part of Reiki except personal music. If we’re doing Reiki, it’s pure Reiki.”* (P6)

##### (d) Support for Reiki Practitioners

In addition to training and modelling the treatment process for new RP, the issue of supporting RP was also generally discussed. One participant spoke of the need to allow a volunteer to say “No” when they didn’t feel comfortable treating someone if it was too emotionally confronting for them.

> *“If they’ve had a family member who’s had a stroke, they may not be comfortable going to see a stroke patient. So they’re free to say, ‘I just can’t go there,’ and we get that.”* (P7)

Another participant spoke of how, as a co-ordinator, she would gather the volunteers together every so often, and they could give each other Reiki and/or just talk about what they are doing as volunteers. She called these gatherings her “Reiki shares.” As discussed earlier, some RP felt supported by other staff who would assist with turning off beeping monitors for part of a session or a hospital that provided a treatment room or massage bed for Reiki sessions. Feeling supported in the challenging environment of a medical setting, especially when unpaid, was important to RP.

#### Theme 3:3 Ethical Considerations for Staff and RP

##### (a) Reiki does not replace medical care, is non-religious, and is not used to diagnose

A number of ethical issues were raised by participants about how Reiki should be explained to staff during staff education sessions and about how Reiki should be delivered by RP. It was emphasized that Reiki ‘adds’ to medical care; it never replaces it. Nor is it ever a cure. However, it may also assist with relaxation, pain relief, and other symptoms.

Participants stressed that this should be made clear to staff during staff training and RP during their training. Emphasizing to staff that Reiki is non-religious was also viewed as important, as was not diagnosing with Reiki.

> *“Reiki practitioners don’t give guarantees… we don’t misrepresent… we don’t diagnose, we don’t prescribe, we’re not a replacement for medical treatment.”* (P2)

> *“I had the experience with someone who told the patient everything they felt. The patient didn’t feel anything. Their expectations weren’t met and they never wanted to do Reiki again… I ended up doing Reiki with that patient with no expectations, just to receive it and it was much better for her.”* (P6)

##### (b) Always offering a choice for Reiki and touch

In a private clinic or some medical centres, the choice to have Reiki is presumed by way of the client having made a booking. But in some medical settings, patients are directly asked whether they would like to receive Reiki. Some also raised the issue of touch. Some said they asked permission and / or delivered Reiki above the person without touching if they preferred.

> *“Celebrate whether they (say) ‘Yes’ or ‘No’, because sometimes they couldn’t say ‘No’ to other treatments. So it was like a gift for them to say ‘No’ or gift for them to receive it.* (P6)

##### (c) Governance, professional membership for RP

This issue of governance and standards was raised by two participants. One participant endorsed allowing only licensed health professionals to deliver Reiki on inpatient wards to ensure they were already familiar with professional standards of practice, while allowing community volunteers in outpatient areas who were still provided with training. Another participant spoke at length about the need for governance through professional membership associations to uphold standards. She believed that more attention should be focused on the code of ethics and conduct for non-registered healthcare workers.

#### Theme 3:4 Promoting Awareness of Reiki

##### (a) Awareness and promotion of Reiki research

All participants felt strongly that spreading awareness of Reiki research and supporting further Reiki research were important to promoting greater acceptance of Reiki in medical settings.

> *“I think more studies, publicity. Everyone who… is a Reiki professional needs to have on their website clear working outlining the benefits of Reiki without misleading.. (they) need to have references to medical studies …clinical trials.”* (P2)

Another participant emphasized the need for Reiki researchers and those funding it to be supported and not suppressed or stigmatized.

> *“There’re people who need to do the research and not be looked at as strange for doing this research, and then there’s people who need to fund the research…. Unless (you have) those two things, people who want to do the research and people who want to fund the research, then it’s not going take off.”* (P8)

##### (b) Acknowledging a spiritual aspect of Reiki

While all participants advocated for Reiki research, a few participants said that for them, it was not just about science. They believed Reiki had a spiritual component that also needed to be acknowledged.

> *“The research will convince people that it’s not just a woo-woo thing… but for me.. it’s more spiritual… (I had) one patient who doesn’t believe in God, ..but after giving Reiki.. he said ‘I’ve never felt a deep, deep peace like this. I’ve never experienced this peace. You know it’s, yeah, it’s beautiful.’ So I think there’s that spiritual aspect as well that needs to be put forward.”* (P5)

##### (c) Management and staff advocating/supporting Reiki

Some participants emphasized the importance of management and staff advocating for and supporting the delivery of Reiki in medical settings through various means. These included not hiding Reiki but championing it.

> *“If people want to start these programs, they have to find their champions. They have to find the people that will always support it. It should never be underground. It should never be like something we’re just kinda hiding… You know what I mean? It should be something accepted. There’s enough I think evidence base out there to support it and there’ll be more obviously.* (P6)

Another participant said that when she introduced herself to patients it made it a lot more difficult it if management hadn’t previously informed patients about what Reiki was and that was non-religious.

##### (d) Providing education about Reiki (internally and externally)

Most participants said one of the most important ways to promote Reiki was through educating staff and patients. The incidence of Reiki education in medical settings amongst participants, however, was highly varied. In some hospitals, there was no education and Reiki appeared hidden or suppressed. At other hospitals where Reiki was more established, Reiki was strongly supported and promoted at all levels of leadership and management. Generally, the more Reiki was promoted through education in the medical setting, the more positive the participants’ felt about providing Reiki at that service, and the more widely Reiki was used and accepted.

> *“The medical providers need more education about complementary therapies and the value of them. I think that’s completely missed in their training…It’s not taking the place of medical treatment at all it’s just providing a way for the patient to have a better experience with their own healing and to be able to release some of the anxiety that goes with what they’re going through.”* (P1)

One participant spoke of how external visiting 4th-year medical students who were taking an integrative therapy elective in their medical school, could come to the hospital and learn about Reiki.

> *“Residents and interns are very, very interested in what this all is and how.. it works?… There’s a course on integrative therapy within the Med school. It’s an elective they can take and they will actually shadow… they’ll go with me for half a day. I’ll do Reiki through them and they’ll be convinced.”* (P7).

##### (e) Teaching Reiki to staff and patients

Teaching Reiki to staff and sometimes patients’ families was also viewed as important in promoting Reiki. Here, not only could Reiki be directly experienced, but it could also be used by staff, patients, and families to heal themselves and others. This in turn promoted awareness of Reiki and its benefits.

##### (f) Unified, consistent explanations of Reiki and Biofield Science

Having clear, consistent explanatory material about Reiki was emphasised when informing or educating staff and patients. One participant said there should also be clear and consistent information about Reiki across promotional materials such as websites and training and educational resources.

> *“Having that wording there, a short spiel that is clear, concise and explains it without the airy fairy BS.”* (P2)

##### (g) Disconnect between intuitive self/ healer and researcher/medical doctor

One participant said that as a medical practitioner, he believed that many other medical practitioners were often disconnected from their spiritual or intuitive selves, while healers tended to be disconnected from their more scientific selves. He suggested that there needs to be more connection between the two.

> *“Most people in the medical field are particularly, you know, lacking in experience… they’re some of the least likely people to be in touch with themselves intuitively, and yet they’re overconfident in their opinions. So there’s a lack of connection amongst the people actually doing the research…And (then), the people who are often healers… they’re not you know, leading researchers… so there’s that disconnect.”* (P8)

## Discussion

### General Findings

This study aimed to qualitatively explore the personal experiences of RP practicing Reiki in medical settings with the hope that this would highlight issues that inform best practice of Reiki in medical settings. Most participants in this study provided Reiki in medical settings free of charge and found it highly rewarding for both them and the patient. They also reported observing and documenting considerable symptom relief and other improvements in the patient. Several RP reported that staff had little or no awareness of Reiki, its research, and sometimes that it was being offered. This was always in medical settings where there was no established Reiki program. Most participants felt supported in their role as a RP, yet despite this, nearly all experienced some negative attitudes towards their practice of Reiki, usually in the form of scepticism. Those who experienced more negative feedback and less support worked in medical settings in Australia where there was no formal educational program for RP and the practice of Reiki was less established.

### The relationship between attitudes and Reiki education

The negative attitudes discussed in these findings align with Hecht’s (2019) findings mentioned earlier in this study. This study highlighted issues raised by licensed mental health professionals (LMHP) who wished to integrate Reiki into their practice but were concerned about judgment from other professionals in their community who may view Reiki as unscientific and be worried about whether the client was open to Reiki (41).

Hecht’s (2019) study also identified that LMHP wanted more education about evidence-based Reiki research for other LMHP and clients. In line with this, participants who worked in medical settings with an established Reiki educational program for RPs and staff reported that Reiki was well received, that they felt supported, and that demand for Reiki was increasing. This suggests that education plays a critical role in determining how well Reiki is accepted and valued. The three longest-running, largest, and well-established Reiki programs were based in the US. These were very well supported by staff and management, with participants experiencing almost no negative attitudes or scepticism. RP in Australian were reasonably well-received, but programs were non-existent, smaller, or less established with little if any education. Generally, these findings suggest more Reiki education for RP and staff promotes less suppression, scepticism, and devaluing of Reiki.

### Feeling unvalued and the issue of remuneration

Another important issue raised by participants was their sense of sometimes feeling unvalued and the question of remuneration. The absence of, or minimal remuneration for RPs in the US and Australia appears to be at odds with the findings discussed earlier of energy healers in the UK (36) which stated 60% of energy healers working in medical centres were being paid. If this discrepancy is correct, it suggests healthcare providers in the UK value the contributions of energy healers more than providers in Australia, and even the US where Reiki appears to be more established in medical settings.

### The Need for Unified Consistent Explanations

In addition to the physical and emotional challenges and the time constraints of working in a medical setting, two other key obstacles were identified for those practicing Reiki in medical settings. The first obstacle was related to a need for consistent, unified, clear language when explaining Reiki, which both staff and patients could relate to. This aligns with the white paper referred to in the introduction, which outlines challenges to the integration of BT (6). It highlighted a need for uniform agreement on accepted terminology, a unified definition, and a clearly defined BF mechanism. Without this, it stated there was confusion among the medical community, educators, and patients. It is not surprising that this need for clear unified explanations would also apply to the integration of Reiki in medical settings. Many health practitioners and allied health staff may simply want to understand what Reiki is and be assured it is safe and helpful, especially when it is paradigmatically at odds with the medical framework in which they practice. Using clear, unified explanations in language health professionals can relate to is important to this goal.

### The Importance of Education

The second key issue for RP in these settings was related to education. The importance of education about Reiki and Biofield Science was perhaps the most important issue to have emerged from this study. It is interesting that in response to question 2: “What are the required conditions for Reiki to be integrated into this medical setting? For example, a required level of research?” No participants mentioned a required level of research for them to practice Reiki in their setting. This may be due to a lack of awareness of Reiki research.

It is also interesting that most RP themselves appeared to have a lack of awareness about the current state of Reiki research, and were unfamiliar with the field of Biofield Science and its related research. These findings are to be expected given the lack of awareness about Reiki research and Biofield Science in general, along with the fact that no participants were researchers in the field. It is also virtually unheard of to see Reiki or Biofield Science research promoted by the media. This finding highlights the limited awareness surrounding the science of Reiki and Biofield Science, and strongly reinforces the need for greater awareness in this area. This is not only critical for the practice of Reiki but also to encourage a broader scientific paradigm shift that incorporates evidence supporting biofield treatments from biological physics, bioelectricity, and biofield science into the current dominant bio-molecular model of health and healing.

The need for education about Biofield Science was also recommended by Guarneri and King (2015) earlier in this paper. They emphasized the need for interprofessional education, which promotes interinstitutional relationships between conventional and CAM institutions. This may include student and faculty exchanges, residency programs, resource modules for BT disciplines incorporated into curriculum materials, and websites to share educational information and resources. It is encouraging that some medical institutions may already be open to this, as one RP described how medical interns had the option to observe RP in a hospital setting as part of their elective in integrative therapy. Other areas highlighted by this paper included the need for a statement of core values that resonates with other disciplines and hosting international, interdisciplinary scientific forums.

### Future Directions: The Need for Interdisciplinary Dialogue

Another issue related to the integration of Reiki into medical settings is a concern that applies to many disciplines, albeit to a lesser extent. This is the issue of what science journalist and author Sally Adee refers to as ‘institutional silos.’ This is the tendency for scientific disciplines to remain insular within themselves, being unaware of and/or resistant to interdisciplinary knowledge and its potential to inform their branch of science (51). Adee (2023), however notes that change has begun. She describes how, in the Department of Biological Engineering at the Massachusetts Institute of Technology (MIT), Cambridge, students can now elect to pursue a PhD in being a Polymath, someone whose knowledge spans many subjects and strives to promote understanding between them. Students learn how to communicate across disciplines by applying concepts to bridge the gaps between them. As participant P8 noted, the emphasis in medicine is on molecular biology, with no understanding or awareness of its relationship to biological physics. A specialist polymath to bridge biofield science and molecular biology may be what is required in the future to assist Reiki’s integration into mainstream health. However, as this can be a barrier between mainstream disciplines (51), overcoming this gap may not occur for some time.

P8 also observed a disconnect between intuitive self / healer types of people and medical practitioner / researcher types and therefore a lack of understanding between the two domains. Encouraging medical practitioners to try Reiki or experiment with their intuitive selves through meditation or other means and encouraging healers to be more aware of empirical research in Biofield Science, an objective mindset, and Reiki research may assist in bridging this gap and promote more understanding between these two domains. Integrating information about Reiki research and even Biofield Science into both medical and RP training may also assist.

### Strengths and Limitations

This study has the following strengths. The qualitative methodology of reflexive TA provided some preliminary foundational insights into what is needed for both RP and other staff when providing Reiki in medical settings. It is also the first study to use a qualitative methodology in exploring the experience of RP in medical settings in Australia and, to the best of the author’s knowledge, internationally. Given the emerging field of Biofield Science and energy therapies, it is important to promote dialogue about the use of these therapies as they become integrated into mainstream settings and to identify what promotes safe best practice. By drawing on the experiences of RPs from the US, where Reiki is more established in medical centres, a deeper awareness was also attained of how Reiki could be better practiced in Australian medical settings.

Regarding limitations, a key aim of this paper was to explore the subjective experience of practicing an alternative BT, Reiki, which is paradigmatically at odds with the setting in which it is practiced – the medical setting. Given this, focusing on the experience of the RP is appropriate as it promotes insight into this experience. Though the questions asked may have partly influenced the topics that emerged. A richer and fairer examination of this issue however, may have been achieved by interviewing other staff members within the medical setting, such as nurses, doctors, and allied health practitioners who are entrenched in the biomedical model. It would be interesting to see to what extent their views align with RP and what concerns they may have about integrating Reiki in their workplace setting. This could be the focus of future research.

Because only a small sample was used across two countries, these results cannot be generalized; instead, they should be used to highlight potentially important issues on this topic. Further research should include a larger sample and/or quantitative methods to broaden our understanding of this area and potentially confirm findings from this study.

### Preliminary Recommendations for Delivering Reiki in Medical Settings

#### 1. Staff Education

It is recommended that all medical, nursing, and allied health staff in medical settings attend an educational session or program about Reiki, delivered in clear, straightforward language, preferably by a health practitioner who uses similar language. This session would explain what Reiki is, research on Reiki’s efficacy, how and where data would be recorded about Reiki’s on-site effectiveness, an opportunity for staff Reiki demonstrations, and an outline of ethical considerations. Ethical information should note that Reiki supports rather than replaces medical care, it is non-religious, it is never used to diagnose, it is only offered with patient consent, and appropriate touch is also only used with patient consent (otherwise it can be done near the body).

#### 2. RP training

It is recommended that RPs complete a training program before working in a medical setting. This training would be additional to a hospital orientation, OHS, and first aid training. It would include information to prepare them for the emotional demands of the medical setting, potential time constraints of competing appointments/visits, and to be mindful of the physical limitations of the setting. It is recommended that RP shadow an existing volunteer for several sessions prior to commencing their work until they are comfortable. Membership with a professional Reiki association is preferred. The ethical considerations outlined to staff should be clearly explained to RP. Simple, clear explanations of Reiki should be agreed upon by RP before they work with patients, and nothing external other than relaxing music should be introduced to the session. The RP should also be supported, and this may include: Reiki shares, debriefing of challenging clients, giving RP a choice to decline to treat a patient, where possible practical assistance (such as turning off the sound from a monitor off during a session, providing a massage bed, or allowing hospital bed adjustments), and the possibility of renumeration or other benefits.

#### 3. Promoting Reiki

Reiki should never be hidden but rather promoted by hospital management and staff through newsletters, flyers, educational materials, and websites that outline what Reiki is, its efficacy to date, what a treatment session entails, and how to book an appointment. A record of Reiki’s on-site effectiveness (pre- and post-session symptom scores) should also be accessible and visible to all staff, for example, on a patient’s daily observation charts and/or in the patient’s file. In this way, all treating medical and nursing practitioners are informed about how the patient responded to Reiki. This data can also be collated and presented at staff education sessions on Reiki. Renumerating RP should be seriously considered by management, particularly where data has been collated showing Reiki has significantly added to therapeutic outcomes. Where appropriate, unified, clear explanations and references about Reiki could also be provided on websites, fliers, or in education sessions about emerging evidence relating to Reiki, and information about Biofield Science, to promote more awareness about this emerging paradigm to staff and the public.

## Conclusions

The practice of Reiki is being widely integrated into some medical settings around the world, particularly in the US and UK. Despite usually being unpaid, RPs report that providing Reiki in these settings was a rich and rewarding experience for both themselves and the patient, and they usually felt supported by hospital staff. However, most RP still experience varying degrees of scepticism and negative attitudes towards their practice from other staff. Key steps towards the successful implementation of Reiki in medical settings include the education of RP to prepare them for medical settings, the education of staff and management about Reiki’s use, efficacy, and ethical considerations, as well as the promotion of Reiki and Biofield Science in these settings. Promotion and education about Reiki and Biofield Science also need to extend to the broader community and always contain clear, unified language and terminology.

## Data Availability

An OSF link to access de-identified transcripts will be provided on request from the first author.

## Acknowledgements

The authors gratefully acknowledge the time of all participants for this study.

## Authors’ contributions

S.Z. conceptualised the study, collected and analysed the data, and completed the draft and final proofs. P.S provided supervision and oversight for the study and proofing of the article.

